# Potential cost-effectiveness of individualized medicine in prodromal Alzheimer’s disease by demographic, clinical and cerebrospinal fluid proteomics factors

**DOI:** 10.1101/2023.01.24.23284478

**Authors:** Ron Handels, Kirsten Wesenhagen, Betty Tijms, Charlotte Teunissen, Pieter Jelle Visser, Linus Jönsson

## Abstract

**INTRODUCTION:** Three distinct pathophysiological subtypes of Alzheimer’s disease (AD) have been identified based on CSF proteomics. We aim to estimate the potential incremental cost-effectiveness ratio of individualized hypothetical AD pharmacological treatment through stratification by these subtypes.

**METHODS:** In a model-based health-economic evaluation usual care was compared to A) hypothetical treatment to in all subtypes and B) hypothetical treatment to those categorized as subtype 1 (high levels of BACE1) based on CSF proteomics; in a population of persons with MCI and positive amyloid and assuming hypothetical treatment efficacy in this subtype 1 only.

**RESULTS:** The potential incremental cost-effectiveness ratio (ICER) was k€36 per quality-adjusted life year (QALY) for strategy A (no subtyping test, hypothetical treatment in all subtypes) as compared to usual care, and k€22 for strategy B (subtyping and hypothetical treatment for subtype 1 only) as compared to usual care. Compared to strategy A, strategy B was dominant, in terms of no difference in QALY and mean cost savings of k€6 per person.

**DISCUSSION:** Given the assumptions in this study, individualized hypothetical AD pharmacological treatment of specific subgroups of AD patients, here based on a proteomics biomarker profile, has the potential to gain health-economic benefits. Future research should focus on retrospective analysis of trial data to generate empirical evidence on treatment effect between CSF proteomics subgroups.

## INTRODUCTION

The global prevalence of dementia has been estimated at 57 million [GBD 2019 Dementia Forecasting Collaborators, 2022] and the corresponding economic impact at US $818 billion [Wimo et al., 2017], reflecting a substantial burden on societies worldwide. In the last decades research has focused on developing diagnostic instruments to identify pathological hallmarks in pre-dementia stages of Alzheimer’s disease (AD) and on developing treatments to delay the onset of dementia for persons living for example with a mild cognitive impairment (MCI) due to AD [Winblad et al., 2016; Cummings et al., 2020].

Various research criteria for AD have been proposed in the last decade to biologically define AD based on biomarkers that indicate the presence of the AD pathological hallmarks, amyloid plaques and tau tangles, in the brain [Jack et al., 2018]. Abnormal biomarker status in non-demented individuals is associated with AD-type dementia at follow-up [Vos et al., 2015]. However, AD is biologically heterogenous, and this complexity is not fully captured by amyloid and tau only. We recently identified 3 AD subtypes with distinct underlying pathophysiological processes based on cerebrospinal fluid (CSF) proteomics [Tijms et al., 2020]. These subtypes may require different treatments. For example, the first subtype showed indications of aberrant neuronal plasticity, which was accompanied by very high levels of BACE1 activity, which is the enzyme involved in amyloidogenic processing of amyloid precursor protein [Vassar et al., 1999] (further referred to as subtype 1). These levels were normal or decreased compared to controls in the other two subtypes. It can be hypothesized that the aberrant neuronal plasticity subtype in particular will benefit from treatments that target APP processing, such as BACE1 inhibitors, whereas this type of treatment may be specifically harmful for individuals with other subtypes. The second subtype was associated with innate immune activation subtype (further referred to as subtype 2), which may potentially benefit from treatments targeting microglia and astrocyte activation. And finally, the third subtype was associated with the blood-brain barrier dysfunction (further referred to as subtype 3), which may benefit from therapies that strengthen the vasculature. These subtypes could be identified in the pre-dementia stages, offering new opportunities for testing secondary prevention strategies.

For an AD treatment in MCI to prove cost-effective it must eventually result in improved health-related quality of life at acceptable required resources. Using CSF-based subtyping to decide upon pharmacological treatment has the potential to prevent treating persons who will not respond to it (i.e., overtreatment). However, inaccuracy in predicting treatment response (e.g., as a result of an imperfect test or imperfect correlation between test outcome and treatment response) could incorrectly withhold individuals from treatment to which they would respond (i.e., undertreatment) [Handels et al., 2015]. To ensure cost-effectiveness, it further requires that a pathological treatment response translates into a timely reduction of clinically relevant outcomes that are associated to health-economic outcomes before death. Early health technology assessment could estimate the health-economic outcomes by simulating the assumed steps of treatment response and clinical outcomes, to support deciding on the further investment in CSF-based subtyping techniques.

We aim to estimate the potential incremental cost-effectiveness ratio of individualized hypothetical AD pharmacological treatment through stratification by CSF-based AD subtypes defined by Tijms et al. [2020] compared to care as usual and compared to no stratification (i.e., same hypothetical treatment in all subtypes), in persons with MCI and abnormal amyloid in CSF in a memory clinic setting.

As estimates of treatment effectiveness by CSF-based subtype are not available we perform an explorative early health technology assessment by means of a health-economic simulation approach in which a scenario of subtype-specific hypothetical pharmacological treatment is evaluated.

## METHODS

A model-based economic evaluation was performed to report time spend in aggregated disease states MCI, mild, moderate and severe dementia, QALYs and costs.

An adjusted societal perspective was chosen that reflected medical, social and informal care.

The simulation ran until the age of 100, which was considered to reflect a lifetime horizon.

A discount rate of 4% for costs and 1.5% for effects was applied.

Based on the predicted lifetime QALYs (around 7 for a person aged 70) the likely applicable willingness to pay (WTP) threshold is €20,000 per QALY gained [Versteegh et al., 2019].

The IPECAD open-source model was chosen for its potential to be adjusted and reflect the target population, setting and strategies (detailed below), as well as its open-source availability. For details on input estimates, see Handels et al. [2022, submitted].

### TARGET POPULATION AND SETTING

The study on the CSF-based subtypes by Tijms et al. [2020] included persons with abnormal CSF amyloid beta1-42 levels, selected from multicenter studies EMIF-AD MBD [Bos et al., 2018] and ADNI (adni.loni.usc.edu). These multicenter studies mainly recruited in a clinical setting and for ADNI sometimes advertised for in the general population. The mean age of the study sample was 68.1 and 74.9 for the selection of individuals from EMIF-AD MBD and ADNI respectively.

The target population of this early health technology assessment was defined as persons with MCI and abnormal CSF amyloid beta1-42, who visited a memory clinic in the Netherlands, aged 70.

### COMPARATORS

A usual care strategy was compared to 2 intervention strategies.

The usual care strategy entailed a standard diagnostic evaluation for persons being referred to a memory clinic for a cognitive complaint. At the memory clinic the syndrome and probable etiology are determined using a standard diagnostic workup including history taking, cognitive, functional, physical, psychiatric and neurological assessment, MRI cerebrum and, if considered relevant, identifying amyloid pathology using CSF or PET. After the diagnosis MCI due to AD (i.e., with abnormal amyloid beta1-42) is established, a tailored treatment plan is developed, which could entail psychosocial support and (annual) retesting cognitive and functional performance until the dementia syndrome and its cause have been established. When dementia is established, treatment with cholinesterase inhibitors could be given and/or supportive formal care could be provided by a general practitioner, case manager or other professional.

Intervention strategy A entailed providing a hypothetical treatment to persons who underwent a lumbar puncture in usual care (i.e., no additional performance of lumbar puncture reflected in this strategy) and were classified as having abnormal amyloid beta1-42 CSF levels. This hypothetical treatment was provided for 5 years. Additional monitoring as compared to the usual care strategy was assumed not required for this intervention strategy. Discontinuation of the hypothetical treatment (e.g., due to side effects) was assumed absent.

Intervention strategy B entailed performing the CSF-based proteomics test to define the subtypes in persons who underwent a lumbar puncture and were classified as abnormal amyloid beta1-42 (same as strategy A). The same hypothetical treatment as in strategy A is provided to the persons with the CSF-subtype test result categorized as ‘aberrant plasticity’ (high levels of BACE1; subtype 1). Absence of additional monitoring and discontinuation was identical to intervention strategy A.

### IDENTIFICATION AND SYNTHESIS OF CLINICAL EFFECTIVENESS DATA

Supplementary table 10 from Tijms et al. [2020] reported 53% of the sample categorized as subtype 1 ((37+66)/193), 24% as subtype ((27+20)/193) and 22% as subtype 3 ((26+17)/193). Furthermore, it reported a hazard ratio of dementia onset of 2.5 for subtype 2 compared to subtype 1, and of 2.1 for subtype 3 compared to subtype 1 based on a Cox regression on the study data [Tijms et al., 2020: supplementary table 15].

A rate for each subtype was calculated by splitting the overall rate into subtype-specific rates in steps (see equation 1). First, the normalized rates for each subtype were estimated 0.62, 1.55 and 1.30 respectively. Second, as the baseline rate (i.e., the number of dementia onset divided by the person-years at risk for dementia) was not reported in the study by Tijms et al. [2020], it was reflected by 0.61 in 3 years obtained from a pooled cohort of data from subjects with IWG-2 criteria based prodromal MCI (defined as any cognitive impairment and ‘abnormal CSF amyloid beta1-42 and tau’ or ‘abnormal amyloid PET scan’) reported by Vos et al. [2015]. This 3-year rate was divided by 3 to obtain a 1-year rate of 0.203. This rate was assumed stable over time and was operationalized by applying a Weibull ‘gamma’ parameter of 1 (i.e., reflecting a stable rate) and calibrated Weibull ‘mu’ parameter of -1.593, to obtain a 1-year rate of 0.203. Third, this baseline rate was multiplied with the normalized rates to obtain the subtype specific rate of 0.13, 0.32 and 0.27. Fourth, they were converted to a transition probability (see equation 2) of 0.12, 0.27 and 0.23 for subtype 1-3 respectively. These were applied to the usual care strategy.

*Equation 1: formula used to split baseline rate to each of the 3 subtypes based on their prevalence and hazard ratio relative to subtype 1*.

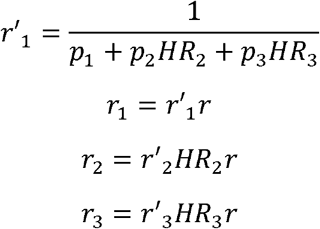

*r’*_*i*_ *= normalized rate subtype i*

*p*_*i*_ *= proportion subtype i*

*HR*_*i*_ *= hazard ration subtype i (subtype 1 as reference)*

*r*_*i*_ *= rate subtype i*

*r = baseline rate*

*Equation 2: formula to calculate transition probability based on rate or rate based on transition probability*.

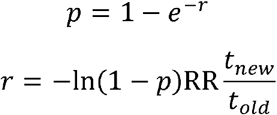

*p = probability*

*r = rate*

*RR = relative risk*

*t*_*new*_ *= targeted time period for the rate (e*.*g*., *1 year, to match the model update cycle)*

*t*_*old*_ *= original time period of the probability (e*.*g*., *2 years, to match the original data source interval period)*

An estimate of treatment effect by subtype from existing treatments was not available from published sources. An effort was made to request trial data for secondary analyses, but data were not shared. Therefore, a hypothetical treatment effect of 20% risk reduction of dementia onset was assumed, similar to previous health-economic analysis evaluating a hypothetical treatment effect using 25% in dementia onset rate [Wimo et al., 2020] or a 20% risk reduction of dementia onset [Green et al., 2019]. Furthermore, due to the lacking data the test outcome was assumed to fully correspond to a treatment response in terms of the clinically relevant outcome of dementia onset (i.e., 100% accurate in predicting treatment response).

This 20% hypothetical treatment effect was assumed observed over all 3 subtypes together (i.e., of any person with abnormal CSF amyloid beta1-42 regardless of subtype), but the effect was assumed to be a result of a treatment response only in persons classified as subtype 1. This was operationalized by multiplying the treatment effect only to the probability of dementia onset in subtype 1. This transition probability was calibrated such that the ratio of total transitions (sum of transitions from subtype 1, 2 and 3 to dementia onset) in the usual care strategy compared to the intervention strategy in the first year was 0.80 (i.e., reflecting a 20% risk reduction). This was obtained if a relative risk of 0.43 in subtype 1 was applied (i.e., a 57% risk reduction). In both intervention strategy A and B, the probability of dementia onset for subtype 1 was multiplied with 0.43.

The hypothetical treatment was assumed of disease-modifying nature (i.e., gained effects will remain but progression will return to its natural progression after treatment is stopped) and to be effective for 5 years, after which the reduced risk of dementia onset was no longer present.

### ESTIMATING UTILITIES, RESOURCES AND COSTS

Utility estimates for MCI and dementia were obtained by pooling the estimates of a selection of sources identified by a systematic review [Landeiro et al., 2020].

Cost estimates for dementia were selected from a systematic review on factors associated to informal care hours and societal costs in dementia [Angeles et al., 2021] and an unpublished systematic review [Landeiro et al., 2018]. Only one study from this review reflected a Dutch setting for mild, moderate, severe dementia for a community and institutionalized living setting [Wubker et al., 2015]. As a cost estimate for MCI in a Dutch setting was not identified, we assumed they were proportional to mild dementia [Leibson et al., 2015].

The costs of the CSF proteomics analysis were assumed €500. This did not include the costs of performing the lumbar puncture (no additional lumbar punctures were performed in the intervention strategies as compared to the usual care strategy).

Costs were transformed to 2020 Euro using EU stats converter [Eurostat, 2021].

### CHOICE OF MODEL

The analyses were performed using the IPECAD open-source decision-analytic model for simulating the effect of a pharmacological intervention in MCI due to AD on disease symptoms and mortality. The model (available as spreadsheet and R code) was chosen for its focus on pharmacological interventions, tackling various model limitations in the field of AD [Nguyen et al., 2018; Gustavsson et al., 2017] and its open-source availability. An individual patient level version of the open-source model was developed to fit the purpose of this study. A basic version of the model is freely available on www.ipecad.org to support transparency and external validation. The version used for this study is available upon reasonable request. A general description of the model can be found from Handels et al. [2022, submitted].

The disease progression part of the IPECAD open-source model was originally developed based on NACC UDC data from the U.S. [Green et al., 2019] with the following details. The starting population represents a selection in this database of persons aged 60-89 with a diagnosis of MCI and AD being reported as the primary or contributing cause of cognitive impairment (i.e., AD indicated as the etiologic diagnosis of the mild cognitive impairment), with AD determined using NINCDS/ADRDA or NIA-AA criteria. Their progression to dementia was modelled by a Weibull function combined with entering in a specific ‘landing’state of dementia severity. For disease progression in dementia a sample of subjects with dementia of AD type (same criteria) was selected and an ordered probit regression was used to predict disease progression in the domains of cognition, function and behavior. In the same sample the probability for institutionalization was estimated based on symptoms in these domains. Mortality was reflected by a combination of increased mortality risk in dementia [Andersen et al., 2010] and assumed not increased in MCI combined with a U.S. age-specific mortality table.

To reflect a Dutch setting the institutionalization rate was calibrated to a Dutch estimate and a Dutch mortality table was used [CBS, 2021].

### ANALYTICAL METHODS

The potential cost-effectiveness of strategy A and strategy B was estimated under the set of base case assumptions. In addition, the outcomes time in each state (MCI, mild, moderate and severe dementia), QALYs and costs were estimated.

A selection of uncertainties was addressed in a univariate sensitivity analysis. Subtype test cost of €2500 was applied. Subtype 1 treatment side effects assumed 0.02 disutility was applied. Treatment response was applied to subtype 2 (microglia and astrocyte activation) and 3 (vasculature protection) instead of 1 (high levels of BACE1).

All data produced in the present study are available upon reasonable request to the authors.

## RESULTS

The potential incremental cost-effectiveness ratio (ICER) was k€36 per QALY for strategy A (no subtyping test, same hypothetical treatment for all) as compared to usual care, and k€22 for strategy B (subtyping and hypothetical treatment for subtype 1 only) as compared to usual care. Strategy B was dominant (in terms of no difference in QALY and mean cost savings of k€6 per person) as compared to strategy A.

Table 1 shows the discounted QALY and cost, and difference for each strategy.

**Table 1:**
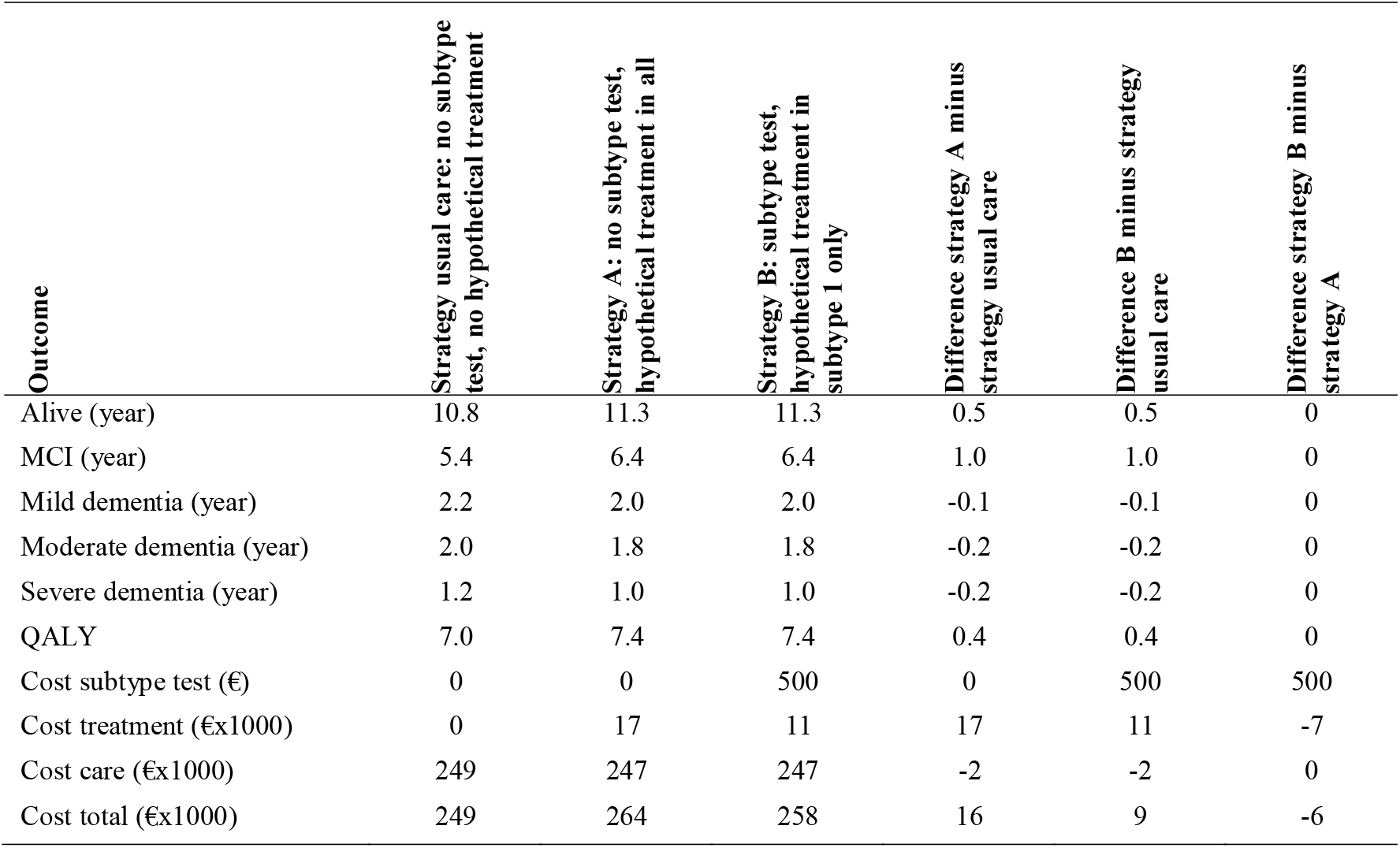
Mean discounted QALY and cost, and difference for each strategy per person.

| Outcome | Strategy usual care: no subtype test, no hypothetical treatment | Strategy A: no subtype test, hypothetical treatment in all | Strategy B: subtype test, hypothetical treatment in subtype 1 only | Difference strategy A minus strategy usual care | Difference B minus strategy usual care | Difference strategy B minus strategy A |
| --- | --- | --- | --- | --- | --- | --- |
| Alive (year) | 10.8 | 11.3 | 11.3 | 0.5 | 0.5 | 0 |
| MCI (year) | 5.4 | 6.4 | 6.4 | 1.0 | 1.0 | 0 |
| Mild dementia (year) | 2.2 | 2.0 | 2.0 | -0.1 | -0.1 | 0 |
| Moderate dementia (year) | 2.0 | 1.8 | 1.8 | -0.2 | -0.2 | 0 |
| Severe dementia (year) | 1.2 | 1.0 | 1.0 | -0.2 | -0.2 | 0 |
| QALY | 7.0 | 7.4 | 7.4 | 0.4 | 0.4 | 0 |
| Cost subtype test (€) | 0 | 0 | 500 | 0 | 500 | 500 |
| Cost treatment (€x1000) | 0 | 17 | 11 | 17 | 11 | -7 |
| Cost care (€x1000) | 249 | 247 | 247 | -2 | -2 | 0 |
| Cost total (€x1000) | 249 | 264 | 258 | 16 | 9 | -6 |

Strategy B (treat subtype 1 only) resulted in 63% less required treatment time to obtain the same effect, corresponding to a mean of €5,750 treatment cost saving per individual. This comes at an additional €500 additional costs per person for performing the subtyping test.

### SENSITIVITY ANALYSIS

When a CSF subtype test costs of €2500 was applied, the potential ICER was k€26 for strategy B (subtyping and hypothetical treatment for subtype 1 only) as compared to usual care. Strategy B was dominant (in terms of no difference in QALY and mean cost savings of k€4 per person) as compared to strategy A.

When a treatment side effect of 0.02 disutility was assumed, the potential ICER was k€43 per QALY for strategy A (no subtyping test, hypothetical treatment for all) as compared to usual care, and k€24 for strategy B (subtyping and hypothetical treatment for subtype 1 only) as compared to usual care. Strategy B was dominant (in terms of mean 0.03 QALY gain and mean cost savings of k€6 per person) as compared to strategy A.

When treatment response was applied to subtype 2 (innate immune activation), the potential ICER was k€77 per QALY for strategy A (no subtyping test, hypothetical treatment for all) as compared to usual care, and k€17 for strategy B (subtyping and hypothetical treatment for subtype 2 only) as compared to usual care. Strategy B was dominant (in terms of no difference in QALY and mean cost savings of k€12 per person) as compared to strategy A.

When treatment response was applied to subtype 3 (blood-brain barrier dysfunction), the potential ICER was k€59 per QALY for strategy A (no subtyping test, hypothetical treatment for all) as compared to usual care, and k€12 for strategy B (subtyping and hypothetical treatment for subtype 3 only) as compared to usual care. Strategy B was dominant (in terms of no difference in QALY and mean cost savings of k€12 per person) as compared to strategy A.

## DISCUSSION

Given the assumptions in this study, individualized hypothetical AD pharmacological treatment (i.e., a combination of testing for AD subtypes and only treating those predicted to respond to it) has the potential to gain health-economic benefits. It showed cost savings and identical QALYs as compared to treat all strategy.

The results of this study are, however, explorative and without an estimate of their uncertainty. This is mainly due to our knowledge lacking evidence on the efficacy of drugs between biologically defined subgroups. The positive health-economic results of this study are therefore not unexpected as the AD subtype test was assumed 100% accurate in predicting clinically relevant outcome of dementia onset. In reality this correlation between subtype test outcome and treatment response is likely to be imperfect, either due to inaccuracy in correspondence to actual underlying subtype (e.g., variation in material handling), due to not fully corresponding to the biological mechanism the treatment is acting upon (e.g., due to mixture of underlying pathology within individuals), or due to heterogeneity in treatment response (e.g., interaction with comorbidity). Nevertheless, confirmation by an additional test that corresponds to some extend to treatment response (i.e., imperfect but reasonable accuracy) in general results in preventing overtreatment at the cost of undertreatment, as has been shown earlier by Handels et al. [2015]. Whether this is cost-effective is mainly determined by the balance between on the one hand cost savings (and prevented side effects) associated to withholding treatment in those correctly re-labelled as non-responders, and on the other hand lost opportunity to gain treatment-generated quality of life in those incorrectly re-labelled as non-responders by the additional test (i.e., prevented overtreatment, and induced undertreatment respectively). Although additional testing trades quality of life for cost savings, this loss might be acceptable as it creates opportunities to generate a potentially higher quality of life elsewhere in the care system, or it might result in access to the AD treatment if affordable only in a subgroup. This is likely to be considered ethically justified if within the subgroup in which treatment is withhold it would not show a significant effect would they have been exposed to it.

Subtype testing is likely a translation of an underlying continuous spectrum into a categorized outcome, of which its cut-off to determine the category can be ranged. This allows to choose a liberal or conservative cut-off, which can be optimized based on its consequential clinically relevant and health-economic outcomes rather than correctness of biological outcomes.

Previous studies have shown the potential of deploying AD related test for treatment allocation to generate health-economic benefits. Handels et al. [2015] indicated a mean headroom of 0.39 QALY and €33,622 savings per person, operationalized as a perfect test with 100% accuracy in identifying AD pathology fully corresponding to a response to a treatment that was assumed already on the market. The QALY gain was related to preventing treatment side effects when treatment was unnecessary (i.e., treating those who would not respond). The cost savings (k€34) were lower compared to this study, probably due to the treatment being applied much longer including during all dementia stages. Sköldunger et al. [2013] found that treatment was more cost-effective in a population enriched with more treatment responders, similar to our findings.

### STRENGTHS & LIMITATIONS

A limitation of the present study is the current lack of evidence on treatment effect within AD subgroups. The results of this study should therefore be considered as an exploration of the potential of subtype testing, supporting discussions on considering investing resources in future research rather than adoption in standard clinical practice.

Various assumptions have been adopted that might influence the study outcomes. A constant hazard was assumed in all 3 subtypes while possibly some of them show accelerating or decelerating rate of dementia onset. A decelerating rate of dementia onset would reduce the headroom for a treatment. However, we expect this not to have a major impact on the incremental effect of subtype testing. The target population is those who have received a CSF test, which was estimated 12% in usual care [Handels et al., submitted] based on a questionnaire among Dutch memory clinics [Gruters et al., 2019]. If, however, CSF subtype testing is cost-effective, it is likely that a larger proportion will receive a CSF test for which its costs should be included in a health-economic evaluation.

An alternative to predicting treatment response using a diagnostic test is to observe treatment response during follow-up monitoring of patient clinical and/or biological outcomes and stop treatment if no response is observed. This allows to save treatment costs, but only after the minimally required treatment duration to allow monitoring a possible response. This scenario should be included in future health technology assessments once both longitudinal data and criteria for determining treatment response are available. As monitoring for side effects is likely part of the required infrastructure when introducing a new treatment, using that same monitoring for determining treatment response could possibly be cost-effective compared to the resources required for additional subtype testing.

Alternative applications of subtype testing such as using it to decide upon further testing using for example amyloid PET or for screening or case-finding in an unselected population have not been considered in this research.

The value of precision diagnostics under the assumption that a treatment only works for 100% or 0% in subpopulation is equal to the avoided amount of unnecessary drug spending and does not require a health-economic simulation model. However, the developed model is ready for the evaluation of future scenarios to include more details and reflect uncertainty.

### RECOMMENDATIONS

For research we urgently advise to share data from completed drug efficacy trials to generate empirical evidence on drug efficacy within CSF-based subtypes. Despite the trials likely not being powered for such subgroup analysis, its results could be used to generate new hypotheses and further support early technology assessment to estimate the potential value of CSF subtyping. If data remains unavailable, a future early health-technology assessment could be performed to explore the value of information from a new trial to that is powered to detect treatment efficacy differences in subgroups.

The results of this study do not provide recommendations for current clinical practice due to the hypothetical situation being reflected in the analyses. Without an existing treatment to be decided upon based on AD subtype, it could nevertheless have an impact on non-medical effects (e.g., impact on emotional, social cognitive or (lifestyle) behavior aspects as a result of knowing ones AD subtype) [Bossuyt et al., 2009] that could be associated to health-economic outcomes. This has not been topic of this study.

### CONCLUSION

This explorative study on the health-economic impact of testing for pathophysiological subtypes of AD in combination with deciding upon a hypothetical treatment showed a potential for being cost-effective, given the assumptions in this study. We believe this supports the recommendation to further invest in research on AD subtype testing and its application in combination with treatment decision making.

## Data Availability

All data produced in the present study are available upon reasonable request to the authors.

http://www.ipecad.org/open-source-model

## ACKNOWLEDGMENT

This work has been supported by ZonMW Memorabel grant program dossier number 70-73305-98-1219.

## AUTHOR CONTRIBUTIONS

RH designed and developed the health-economic model, generated the results and drafted the report. LJ designed the health-economic model. KW performed statistical analyses to generate inputs for the health-economic model. All authors interpreted the results, revised the report and approved the final version.

## COMPETING INTERESTS

**Ron Handels** reports the following related to this study: grant (paid to department) from ZonMw (NL public funding; 2017-2024 dossier number 70-73305-98-1219). Ron Handels reports the following in the past 36 months outside this study: Funding (paid to department) from Karolinska Institutet via affiliation, related to projects: SNAC (Sweden public funding 2016-2018), MIND-AD (public-private EU JPND grant 2017-2018), PRODEMOS (public EU H2020 2019-2023), SveDem (Sweden public-private collaboration 2019-2020), EUROFINGERS (public-private EU JPND; 2020-2023); funding (paid to department) from Radboud University Nijmegen, related to project: AI-MIND (EU H2020 public funding 2021-2023); grants (paid to department) from RECAGE H2020 (EU public funding; 2018-2022); grants (paid to department) from various ZonMw and NWO projects (NL public funding; 2017-20284); grants (paid to department) from patient association Alzheimer Nederland (NL fellowship; 2017-2019; WE.15-2016-09); grants (paid to department) from ROADMAP (IMI2; public-private collaboration; 2016-2019); consulting fees (paid to department) from Lilly Nederland B.V. (advisory; 2022-2023); consulting fees (paid to department) from institute for Medical Technology Assessment (advisory; 2021; content initiated by Biogen); consulting fees (paid to department) from Biogen Netherlands BV (advisory; 2021); consulting fees (paid to department) from Biogen MA Inc. (advisory; 2020); consulting fees (paid to department) from Eisai Inc. (advisory; 2019).

**Linus Jönsson** reports the following related to this study: none. Linus Jönsson reports the following in the past 36 months outside this study: employment at H.Lundbeck (ended 2021).

**Charlotte Teunissen** is supported by the European Commission (Marie Curie International Training Network, grant agreement No 860197 (MIRIADE), Innovative Medicines Initiatives 3TR (Horizon 2020, grant no 831434) EPND (IMI 2 Joint Undertaking (JU) under grant agreement No. 101034344grant no) and JPND (bPRIDE), National MS Society (Progressive MS alliance) and Health Holland, the Dutch Research Council (ZonMW), Alzheimer Drug Discovery Foundation, The Selfridges Group Foundation, Alzheimer Netherlands, Alzheimer Association. Charlotte Teunissen is recipient of ABOARD, which is a public-private partnership receiving funding from ZonMW (#73305095007) and Health∼Holland, Topsector Life Sciences & Health (PPP-allowance; #LSHM20106). ABOARD also receives funding from Edwin Bouw Fonds and Gieskes-Strijbisfonds. Charlotte Teunissen has a collaboration contract with ADx Neurosciences, Quanterix and Eli Lilly, performed contract research or received grants from AC-Immune, Axon Neurosciences, Bioconnect, Bioorchestra, Brainstorm Therapeutics, Celgene, EIP Pharma, Eisai, Grifols, Novo Nordisk, PeopleBio, Roche, Toyama, Vivoryon. She serves on editorial boards of Medidact Neurologie/Springer, Alzheimer Research and Therapy, Neurology: Neuroimmunology & Neuroinflammation, and is editor of a Neuromethods book Springer.

**Betty Tijms** and **Pieter Jelle Visser** are the inventors of the CSF proteomic profile definition of Alzheimer’s disease sub-types and have a patent pending (#19165795.6; Applicant: Stichting VUmc).

**Kirsten Wesenhagen** declares no competing interests with the content of this article.

